# A protocol for MIndfulness-based Neurofeedback to augment DBT psychotherapy for adults with Borderline Personality Disorder (MIND-BPD)

**DOI:** 10.1101/2025.11.18.25340531

**Authors:** Katherine G. Jones, Marlee M. Vandewouw, Jitendra Awasthi, Alexandra A. Alario, Clemens Bauer, Beth Brewer, Nicole Campbell, Dominic Denning, Keara D. Greene, Jude Hammoud, Oliver Hinds, Sarah Huffman, Connie Maerker, Sarah Paprotna, Maolin Qiu, Benjamin Swinchoski, Anna Taylor, Elinor Waite, Paul Wighton, Maya Whaley, Jillian Papa, Katherine Dixon Gordon, Michelle Hampson, Susan Whitfield-Gabrieli, Sarah K. Fineberg

## Abstract

Borderline personality disorder (BPD) is a severe psychiatric condition associated with high rates of suicide and poor interpersonal functioning. There are no FDA-approved medications, and evidence-based psychotherapies such as dialectical behavior therapy (DBT) are difficult to access and vulnerable to patient dropout. Novel treatment directions are urgently needed. Mindfulness is the core skill in DBT. Our team has developed a mindfulness-based real-time neurofeedback (mbNF) paradigm where individuals learn to reduce default mode network (DMN) versus control network activation in order to increase present-moment awareness. Here, we describe the study protocol for the MIndfulness-based Neurofeedback to Augment DBT Psychotherapy for Borderline Personality Disorder (MIND-BPD) trial. Participants with BPD (N = 52) enrolled in the study will be randomly assigned (1:1 ratio) to receive one session of either real or sham mbNF. Following neurofeedback, all participants will be enrolled in a 6-month DBT psychotherapy group. Supported by a R61/R33 grant from the NIH, the primary outcome of the R61 phase of this trial is change in DMN connectivity between pre- and post-NF resting-state scans, as defined by increased within-network connectivity between the medial prefrontal cortex (mPFC) and posterior cingulate cortex (PCC), and increased connectivity between the mPFC and dorsolateral prefrontal coftex (dlPFC). Change in self-reported mindfulness between pre-and post-NF is a key secondary outcome. This study is registered in the US Clinical Trials Registry (NCT06446765).

## Introduction

Borderline personality disorder (BPD) is a common and debilitating psychiatric condition. It affects ∼2% of the population (1) and is associated with emotional lability, social difficulties, unstable sense of self and risky behavior. These experiences can be distressing, and lead to impairment in personal and work relationships (2). In addition, people with BPD have high rates of suicidality: 5-10% complete suicide (1, 3). The current treatment landscape is insufficient. There is no well-evidenced or guideline-recommended medication; evidence-based psychotherapies are not widely available and have high rates of dropout (4, 5), and many patients are left with residual symptoms. People do get better from BPD, showing decreased symptoms over time, but as many as 40% of people with BPD do not achieve functional recovery (6). More effective and efficient treatment strategies are urgently needed.

Dialectical behavior therapy (DBT) is an evidence-based treatment for BPD. The originally tested implementation of DBT is a 12-month program with individual and group therapy, as-needed phone coaching to support use of therapy skills between sessions, and a consultation team to support the therapists. Group sessions focus on four topics: mindfulness, distress tolerance, emotion regulation, and interpersonal effectiveness (7, 8). While DBT studies have investigated this comprehensive model, early evidence supports the efficacy of skills training group (DBTsg) to reduce symptoms without simultaneous individual DBT therapy (9, 10).

Mindfulness — the intentional practice of non-judgmental acknowledgement and awareness of the present moment — is the core skill in DBT (11). Deficits in mindfulness are consistently linked to BPD severity (12–15), and mindfulness training improves BPD symptoms (16). Moreover, compared with other DBT modules, mindfulness has been associated with decreased impulsivity (17) and decreased BPD symptoms overall (18). For DBT patients, mindfulness theoretically facilitates emotional awareness, emotion regulation, and effective action. For this reason, mindfulness training precedes each of the DBTsg modules, preparing patients to learn from skills-based therapy (7).

At the level of neural circuits, mindfulness is inversely associated with functional connectivity in the default-mode network (DMN). The DMN is active during self-referential and internally directed thought, and is anti-correlated with network active during task engagement (task-positive networks), such as the central executive network (CEN) (19). Increased mindfulness is associated with decreased DMN connectivity (20). A meta-analysis of seven large studies in adults with BPD found hyperconnectivity between DMN hubs medial prefrontal cortex (mPFC) and posterior cingulate cortex (PCC) (19), pointing to aberrant DMN connectivity in BPD. Therefore, we hypothesize that mindfulness training can decrease DMN hyperconnectivity and improve symptoms in BPD.

Neurofeedback is a technique that trains individuals to modulate their own brain activity by providing real-time feedback on neural signals using modalities such functional magnetic resonance imaging (fMRI). The high spatial resolution of fMRI allows researchers to deliver real-time neural feedback to probe and strengthen specific brain regions or circuits associated with behaviors of interest; these neural changes can be empirically verified by post-training fMRI scans. fMRI neurofeedback has shown utility as an intervention in both clinical and non-clinical populations, and early clinical trials have reported promising results across a range of disorders and symptoms, including depression (21, 22), auditory hallucinations (23), contamination anxiety (24), Parkinson’s disease (25), phobia (26), and Tourette’s syndrome (27). Early data also suggest therapeutic effects of fMRI neurofeedback paradigms in BPD (28, 29).

Our group has developed an innovative fMRI neurofeedback paradigm to augment mindfulness practice: mindfulness-based neurofeedback (mbNF) (21, 23). Here, participants see a visual display of the difference between real-time DMN and CEN activation levels and use mindfulness as a strategy to volitionally regulate this difference (21). In non-BPD clinical samples, mbNF has been shown to reduce within-DMN connectivity in adolescents with a history of anxiety and/or depression (21), and increase DMN-CEN anticorrelations in adult patients with schizophrenia (23). In addition, both populations saw changes in symptom scores: increased state mindfulness in adolescents with affective disorder history (21) and lower auditory hallucination symptoms in schizophrenia (21, 23).

The present study addresses the urgent need to optimize treatments for BPD and extends this to the evaluation of a novel intervention. In this manuscript, we describe the protocol for a trial that combines mbNF with DBTsg in patients with BPD.

## Methods

### Study Overview

The MIndfulness-based Neurofeedback to augment DBT psychotherapy for adults with Borderline Personality Disorder (MIND-BPD) study is a multi-site double-blind randomized controlled trial in adults with BPD. Treatment-seeking adults with BPD are randomly assigned (1:1 ratio) to one session of mbNF or sham neurofeedback. All participants proceed to 20 weeks of DBTsg after the neurofeedback session. Study participants and clinical raters are blinded to treatment allocation. The project is approved by the Yale Institutional Review Board (HIC#2000037582, first approved June 17, 2024) and is a collaboration between three academic centers (Yale School of Medicine, Massachusetts General Hospital (MGH), and University of Massachusetts Amherst (U Mass)) and an independent contractor (Jillian Papa of Eval for Impact). The Yale site serves as the coordinating center and conducts all recruitment and clinical assessments. fMRI scanning and data analysis occur at both Yale and MGH. Psychotherapy is conducted remotely by the team at U Mass. Program evaluation and lived experience input is overseen by Jillian Papa. An independent Data Safety and Monitoring Board oversees the conduct of the study. The trial is registered in the US Clinical Trials Registry (NCT06446765). The study is funded by the National Institute of Mental Health (R61MH135009) under the R61/R33 mechanism, which affords two years for the first phase of work to establish target engagement (the R61 phase). Then, a data-driven go/no-go decision will inform whether or not the project can progress to the R33 three-year phase to establish clinical efficacy. This manuscript describes the R61 phase activities in detail (recruitment through neurofeedback day) and briefly outlines the post-neurofeedback day psychotherapy and post-psychotherapy follow-up activities that are the intended focus of the R33 phase. Recruitment began in March 2025 and is expected to be complete in August 2025, with all data for the R61 primary outcomes collected by September 2025 and results reported shortly thereafter (September-October 2025).

### Recruitment

Study participants are recruited from the community in Connecticut and Massachusetts due to geographic access to the scan sites and limits of clinician licensure for DBTsg. Advertisements are disseminated on social media and through clinical services. All study activities are conducted in line with a protocol approved by the Yale Institutional Review Board serving as single IRB for the study.

### Inclusion/Exclusion Criteria

Participants complete a consent process including verbal review of study rationale, procedures, risks/benefits, privacy practices, and compensation. Participants who provide written consent are scheduled for a series of in-person or video conference meetings for full study eligibility assessment (see inclusion/exclusion criteria in **Table 1**). Demographics, treatment history, and history of developmental, learning, and medical problems are assessed with interviews and surveys. Careful assessment of MRI safety is conducted using a detailed standard safety questionnaire at baseline and again on scan day. Participants are assessed for adequate English skills to complete study interviews and surveys and to respond to instructions during MRIs.

**Table 1:**
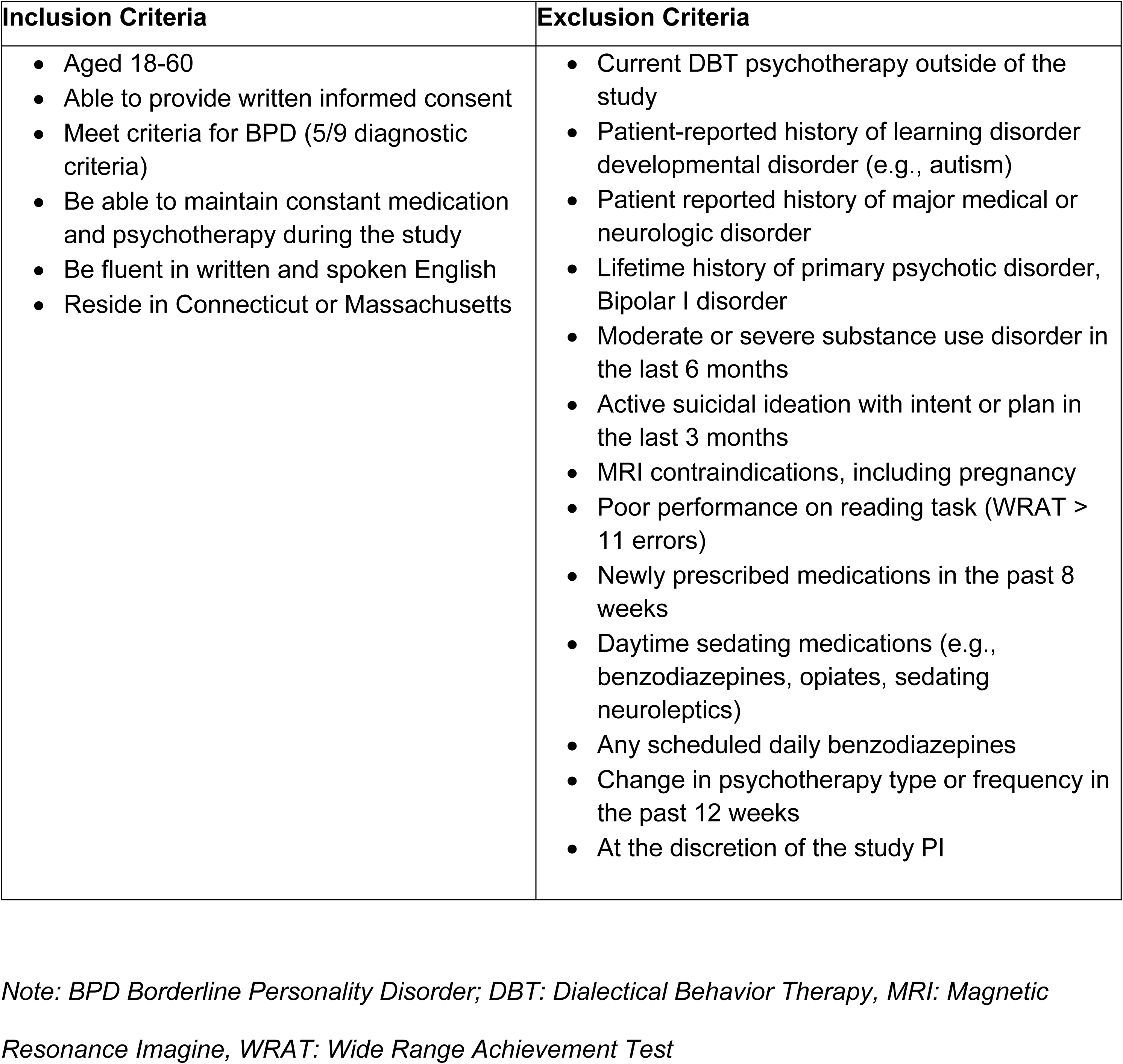
Study Inclusion and Exclusion Criteria.

| Inclusion Criteria | Exclusion Criteria |
| --- | --- |
| • Aged 18-60<br>• Able to provide written informed consent<br>• Meet criteria for BPD (5/9 diagnostic criteria)<br>• Be able to maintain constant medication and psychotherapy during the study<br>• Be fluent in written and spoken English<br>• Reside in Connecticut or Massachusetts | • Current DBT psychotherapy outside of the study<br>• Patient-reported history of learning disorder developmental disorder (e.g., autism)<br>• Patient reported history of major medical or neurologic disorder<br>• Lifetime history of primary psychotic disorder, Bipolar I disorder<br>• Moderate or severe substance use disorder in the last 6 months<br>• Active suicidal ideation with intent or plan in the last 3 months<br>• MRI contraindications, including pregnancy<br>• Poor performance on reading task (WRAT > 11 errors)<br>• Newly prescribed medications in the past 8 weeks<br>• Daytime sedating medications (e.g., benzodiazepines, opiates, sedating neuroleptics)<br>• Any scheduled daily benzodiazepines<br>• Change in psychotherapy type or frequency in the past 12 weeks<br>• At the discretion of the study PI |

*Note: BPD Borderline Personality Disorder; DBT: Dialectical Behavior Therapy, MRI: Magnetic Resonance Image, WRAT: Wide Range Achievement Test*

Trained interviewers conduct clinical interviews using semi-structured instruments. For BPD assessment, we use the Diagnostic Interview for Personality Disorders BPD questions (30). If an individual without a prior BPD diagnosis is found to meet criteria, a licensed clinician facilitates a collaborative diagnostic disclosure. To assess for primary psychotic disorder, Bipolar I disorder, and alcohol and substance use disorders, we use the Structured Clinical Interview for DSM-5 for general psychopathology (SCID5) (31). For assessment of suicidal ideation and behavior, we use the Columbia Suicide Severity Rating Scale (C-SSRS) (32).

Polypharmacy is common in BPD (33–35); this study allows most medications to increase the generalizability of results to a real-world population. All medications are documented and will be considered in analyses.

### Study Design

This study is a prospective, randomized, two-arm double-blinded clinical trial of active versus control neurofeedback to augment evidence-based psychotherapy (DBTsg) for the treatment of adults with BPD. Study activities are detailed in a SPIRIT diagram (**Figure 1**). Participants in both groups receive 20 weeks of remote (video-conferenced) DBTsg (one 2-hour session each week). The protocol proceeds through four stages: I — Enrollment (eligibility assessment and baseline measures), II — Intervention (personalized brain mapping and neurofeedback), III — DBT skills group with intermittent assessments, and IV — Post-treatment follow-up. Stage I usually occurs over two weeks to two months. Stage II is one day (the outcomes for the R61 phase of the project are measured during this stage). Stage III includes 20 consecutive weekly DBTsg sessions (adjusted for state or national holidays). During stage III, clinical assessments are administered every 5 weeks coinciding with the start of each of the 4 DBT modules. Stage IV lasts two months, with two monthly clinical assessments and one follow-up fMRI scan in post-therapy month two.

**Fig 1.**
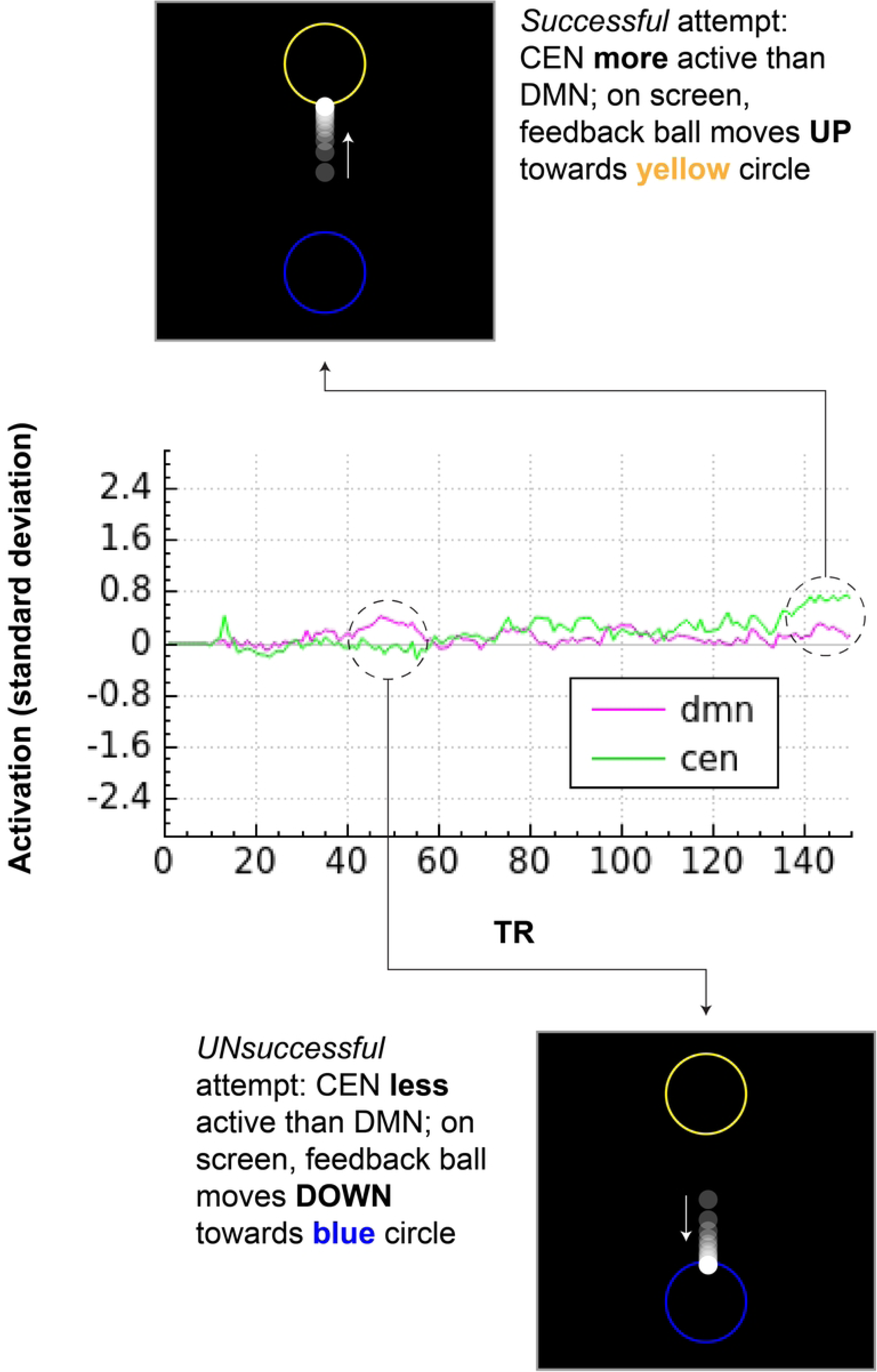
Participant timeline: Schedule of enrollment, interventions, and assessments. Note: WRAT: Wide Range Achievement Test, DIPD-BPD: Diagnostic Interview for Borderline Personality Disorder BPD section, SCID: Structured Clinical Interview for DSM-5 research version, CSSRS: Columbia Suicide Severity Rating Scale, mbNF: mindfulness-based Neurofeedback, EHI: Edinburgh Handedness Inventory, MAAS: Mindfulness Attention Awareness Scale, DSM-5: Diagnostic Statistical Manual, 5^th^ Edition, PHQ9: Patient Health Questionnaire 9-item version, GAD-7: Generalized Anxiety Disorder scale 7-item version, DES: Dissociative experiences scale, LEC: Life Experiences Checklist, PCL-5: PTSD Checklist for DSM-5, UWRAP: University of Washington Risk Assessment Protocol, SMS: State Mindfulness Scale, BSL-23: Borderline Symptom List 23-item version, IMI: Intrinsic Motivation Scale, SAS: Social Adjustment Scale Self-report, DERS: Difficulties in Emotion Regulation Scale 18-item version, DIRE: Difficulty in Interpersonal Emotion Regulation, PIES: Perceived Invalidation of Emotions Scale, SoA: Sense of Agency scale, RAS: Recovery Assessment Scale; SITBI-R: Self-injurious thoughts and behaviors inventory revised, WHODAS: World Health Organization Disability Assessment Scale

### Study Interventions

#### Randomization and blinding

Participants who complete the MRI localizer session (described below) are randomized within-site to receive either active or control neurofeedback (see details in *Control condition* section below) using a predetermined randomization schedule prepared by and known only to the site neuroimaging staff. Clinical staff and assessment raters remain blind to condition assignment. Each site has 1:1 randomization, with the constraint that the number of active NF cases is always greater than or equal to the number of control NF cases (necessary to ensure each control participant has a prior active NF participant to be matched to), and also the constraint that the number of active NF cases does not exceed the number of control NF cases by more than three (to keep the two groups reasonably balanced over time). If a participant is randomized but does not complete the NF MRI session, they are replaced (the next participant uses their randomization number) and they do not continue with study activities. Participants are debriefed at the end of the study, and at that time their group assignment is disclosed.

#### Mindfulness-based neurofeedback (mbNF)

Participants who meet eligibility criteria following intake and assessments complete a neuroimaging session at either Yale or the Martinos Center for Biomedical Imaging at MGH (**Figure 2**). The session begins with baseline structural, diffusion, and resting-state functional MRI acquisitions, which serve as the localizer for generating individualized network maps used in neurofeedback. After scanning, participants receive guided instruction in mindful describing (see details below), which they are encouraged to use in the MRI during the subsequent neurofeedback session.

**Fig. 2.**
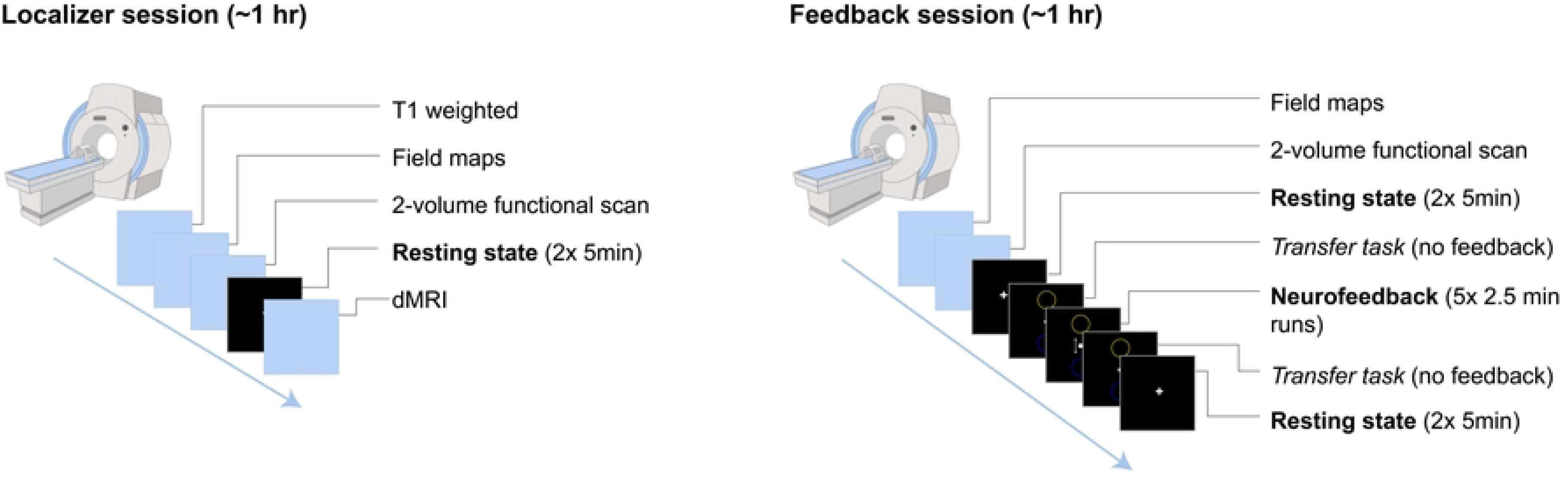
Mindfulness-based Neurofeedback Procedure Note: dMRI – diffusion MRI

#### MRI data acquisition

All MRI data will be collected on 3T Siemens scanners with a 64-channel head coil: a Vida system at Yale and a Skyra system at MGH. Sequence parameters have been harmonized across both sites to the extent permitted by hardware and software constraints; parameters for Yale are presented in **Table 2**. Imaging includes a high-resolution T1-weighted structural MRI (sMRI) and diffusion-weighted MRI (dMRI) sequences. Functional imaging (fMRI) comprises both resting-state and NF sequences and includes two images with opposite phase encoding for offline fieldmap correction.

**Table 2:** MRI acquisition parameters.

| Parameter | sMRI | dMRI | fMRI |
| --- | --- | --- | --- |
| Sequence | MPRAGE | EPSE | EPI |
| TR (ms) | 2300 | 4000 | 1200 |
| TE (ms) | 2.26 | 110 | 30 |
| TI (ms) | 900 | – | – |
| FA (°) | 8 | 78 | 90 |
| Matrix | 256×256 | 140×140 | 98×98 |
| Slices | 192 | 92 | 75 |
| FOV | 256×256 | 210×210 | 200×200 |
| FOV phase (%) | 100 | 100 | 100 |
| Resolution (mm) | 1×1×1 | 1.5×1.5×1.5 | 2×2×2 |
| Parallel acceleration factor | 2× | None | 2× |
| Diffusion directions | – | 198 | – |
| *b*-values (s/mm<sup>2</sup>) | – | 0, 1500, 3000 | – |

*Note: MRI: Magnetic resonance imaging; sMRI: structural MRI; dMRI: diffusion MRI; fMRI: functional MRI; TR: repetition time; TE: echo time; TI: inversion time; FA: flip angle; FOV: field of view; MPRAGE: magnetization-prepared rapid gradient echo; EPSE: echo planar spin echo; EPI: echo planar imaging*

#### Localizer session and generation of personalized network maps

At the start of the scan visit, participants complete a localizer session consisting of sMRI, dMRI, and 10 minutes of resting-state fMRI. These scans serve dual purposes: characterizing baseline anatomy and functional connectivity and generating individualized network maps to guide NF. This ensures that the feedback signal reflects each participant’s unique functional architecture.

While participants complete the mindful describing training outside the scanner, the resting-state fMRI data are preprocessed using FMRIB’s Software Library (36). Preprocessing steps include motion correction, brain extraction, co-registration to standard space, spatial smoothing (5mm full width at half maximum [FWHM]), and high-pass temporal filtering (0.01Hz). Independent component analysis is applied to extract approximately 30 components. These components are spatially correlated with canonical network templates from the Yeo et al. 2011 parcellation to identify participant-specific maps of the DMN and CEN (37). For each network, the component with the highest correlation is selected, thresholded to retain the top 10% of voxels by loading strength, binarized, and registered to the participant’s native space.

#### Mindful Describing training

After the localizer session, participants come out of the scanner for mental noting training (here termed “mindful describing” to cohere with DBT terminology). This semi-structured mindfulness training protocol, established in our prior work (23), is administered by trained research staff in a 45-minute in-person lesson. This lesson describes how to do the practice and introduces a behavioral and neural rationale for the approach, including introducing the goal of activating the CEN versus the DMN. The mindful describing practice focuses on present-moment awareness; participants learn to mentally label present-moment sensations (e.g. seeing, hearing, feeling).

They are discouraged from focusing on thinking instead of perception, and from focusing on breath, which could confound fMRI data during the mbNF acquisition. Each participant selects an “anchor,” a part of their own body they can return to focusing on if they lose track of the practice. Each participant is trained until they and their trainer feel they are competent to use mindful describing in the scanner. This is assessed after several practice rounds: practicing mindful describing aloud to study staff, silently in their heads, and silently while hearing simulated MRI background noise. Participants are encouraged to use the skill during the neurofeedback training session and to consider during and after training how they could use mindful describing to manage distress in daily life.

#### Neurofeedback: Paradigm

Following mindful describing training, participants return to the scanner for the neurofeedback session (**Figure 1**). A brief functional scan is first acquired to co-register the participant-specific network maps to their current position in the scanner. This is followed by 10 minutes of resting-state to establish post-training, pre-neurofeedback baseline connectivity.

Participants then complete one 2.5-minute “transfer” run prior to neurofeedback, in which the neurofeedback display is visible but static and participants view the interface without receiving real-time feedback. This allows for the assessment of whether the learned regulation strategies persist in the absence of feedback. The core neurofeedback protocol consists of five 2.5-minute runs of mbNF. During these runs, participants practice mindful describing while receiving visual feedback based on the difference in activation between the CEN and DMN, guiding participants towards decreased DMN activity and increased CEN activity. (see *Neurofeedback: Implementation* for details). The session concludes with a second transfer task and 10-minute resting-state acquisition to assess post-mbNF connectivity.

#### Neurofeedback: Control condition

Determining a control condition for non-medication interventions can be complex, as there is often no readily apparent placebo; neurofeedback is no different (38). This study employs a yoked-sham control condition. Participants in both the control and active NF groups complete the mindfulness training and undergo similar MRI scan procedures. The primary difference is that the control NF participants do not receive feedback based on their own brain activity, but instead view feedback derived from a previously acquired active mbNF session from another participant. Thus, the visual display in the control group is independent of their own neural activity, ensuring that participants across groups are exposed to equivalent stimuli.

The yoked sham control approach enables participant blinding, provides a similarly engaging feedback experience, and balances the amount of positive feedback received across groups. Specifically, each control participant receives feedback indicating they are achieving the same amount of success as the active NF participant to whom they are yoked. Perceived success can influence motivation, as individuals are more likely to remain engaged when they feel they are doing well. It may also amplify placebo effects, as seeing apparent success in regulating mindfulness-related brain patterns can enhance expectations and perceived efficacy. Therefore, controlling for perceived success is critical.

An alternate control condition used in some studies involves training participants to regulate a brain region unrelated to their symptoms (e.g., from the somatomotor network). However, this approach has the disadvantage that the ease of modulating this region may differ between the control and active NF groups, as some brain areas are more easily regulated than others. This can lead to unequal perceptions of success on the task, which in turn may introduce differences in motivation and placebo effects.

One concern common to all sham-controlled designs is that participants in the control group may notice that the feedback does not correspond with their emotional state, potentially compromising the blinding. For this reason, participants are asked at the end of the NF session to report their beliefs about group assignment (i.e., which intervention they think they received, and their confidence in this belief).

Overall, by providing maximally similar procedures across active and control groups, we aim to account for potential mindfulness practice effects (e.g., learning, utilization) and non-specific scanner effects.

#### Neurofeedback: Implementation

Real-time NF is implemented using the multivariate and univariate real-time functional imaging (MURFI) (39) software in conjunction with PsychoPy. A high-performance computer connected via Ethernet to both the MRI scanner and stimulus presentation system enables real-time data transfer and display. As functional volumes are acquired, MURFI computes the NF signal, which is transmitted to PsychoPy for dynamic display updates.

The NF signal is derived from a positive diametric activity (PDA) metric, defined as the difference in activation between the CEN and the DMN. For each acquired volume, an incremental general linear model estimates expected signal based on nuisance regressors, including six head motion parameters and a linear trend. The residual signal, representing neural activation, is computed by subtracting the nuisance signal from the observed fMRI intensity and *z*-scored relative to a 30-second baseline. Network activation is calculated as the weighted mean of residuals across voxels, with weights inversely proportional to baseline noise. The PDA metric is then computed as the difference between CEN and DMN activation and used to drive the feedback display.

During neurofeedback runs, participants view a visual interface consisting of a centrally positioned white dot and two flanking circles (**Figure 3**). The dot represents the current neural activity, and its position updates in real time based on the PDA metric. When CEN activation exceeds DMN activation, the dot moves upward towards the top circle; when DMN activation is dominant, the dot moves downward toward the bottom circle. This intuitive display guides participants toward decreased DMN activity while practicing mindful describing.

**Fig. 3:** Participant view and corresponding activity during neurofeedback Note: CEN: central executive network; DMN: default mode network

#### Dialectical Behavior Therapy skills group (DBTsg)

Within ten days of completing NF, participants begin DBTsg. The groups are led by trained Masters- and PhD-level staff trainees from the UMass Psychological Services Center who have completed at least the equivalent of foundational training in DBT. Each group is led by two leaders, consistent with the DBT manual (7), who are supervised by a licensed clinical psychologist (KDG) and attend a weekly DBT consultation team. The groups follow the 20-week curriculum developed by McMain et al., which improves emotion regulation, distress tolerance, anger, and suicidal/self-harming behaviors (40). The group curriculum draws on the standard DBT manual, and teaches skills across distress tolerance, interpersonal effectiveness, dialectics, and emotion regulation. Each of these modules is preceded by one session focused on mindfulness. DBTsg runs continuously, with rolling admission: New participants join at the beginning of each module, and graduate once they have completed all sessions. In this way, we enroll participants continuously through the life of the project because DBTsg start dates occur approximately every five weeks.

### Outcome measures

#### fMRI outcome measures

The primary outcome for the R61 study is the change in two features of DMN connectivity between pre- and post-NF resting-state scans: (1) within-network connectivity between the medial prefrontal cortex (mPFC) and posterior cingulate cortex (PCC), and (2) DMN-CEN anticorrelation, defined as connectivity between the mPFC and dorsolateral prefrontal cortex (dlPFC).

The resting-state data will be processed using fMRIPrep (41) and the CONN toolbox (42). Preprocessing steps include slice timing correction, motion correction, susceptibility distortion correction using fieldmap-based unwarping, co-registration to the T1-weighted anatomical image, normalization to standard space, and spatial smoothing (4mm FWHM). Denoising will include regressors for the six motion parameters and their temporal derivatives, the top five components capturing signal from the white matter and cerebrospinal fluid (43), outlier volumes defined by a framewise displacement >0.5 or standardized DVARS>1.5, and a linear trend term. Bandpass filtering will be applied to retain frequencies between 0.008 and 0.09Hz.

Regions of interest (ROIs) for the mPFC, PCC, and dlPFC will be defined based on 10mm spheres centered on established coordinates (44). Mean timeseries will be extracted from each ROI, and Pearson correlation coefficients will be computed between the mPFC and PCC (within-DMN connectivity), and between the mPFC and dLPFC (DMN-CEN anticorrelation). We will also explore generating participant-specific ROIs by identifying the peak voxel within each ROI, and constructing a 10mm radius sphere centered on that peak.

#### Patient-reported measures

The secondary outcome measure for the R61 study is change in state mindfulness from before to after mbNF on scan day. The State Mindfulness Scale (SMS) is a 21-item self-report measure that quantifies current-moment mindfulness of body and mind (45). Change in mindfulness in the study will also be measured by querying self-reported use of mindfulness skills in daily life at baseline and subsequent assessment timepoints.

Several scales are used to track clinical and psychological change in the study, though these are not the focus of the R61 phase of the project. The Borderline Symptom List (BSL-23) is a 23-item scale that measures BPD symptom severity over the past one week (46). The Beck Depression Inventory (BDI) is a 21-item scale that measures depression severity over the past two weeks (47–49). We also track DBTsg group attendance and use of mental healthcare resources outside of the study to support exploratory analyses.

Further psychological measures are collected to support exploration of change in shame (50), social functioning (51), emotion regulation (52–54), sense of agency (55, 56), and episodic future thinking (57) over the course of the study.

Additionally, several measures are collected at baseline to contribute to description of the sample and for exploratory analyses of heterogeneity of treatment effects, including demographics, measures of handedness, quality of life, and psychiatric symptoms (see details in **Figure 1**).

#### Lived experience-led program evaluation

This study incorporates assessment of participant experience at multiple time points to support the development of an intervention that is well-tolerated by the target population.

To evaluate tolerability, study staff assess both physical and emotional adverse events before and after neurofeedback sessions. Prior to neurofeedback, participants are interviewed about their expectations about the intervention. Following neurofeedback, they complete the Intrinsic Motivation Inventory (IMI) — which measures motivation, likelihood of future participation, and enjoyment (58) — and participate in a semi-structured qualitative interview exploring their experience in the scanner and strategies used to maintain focus.

A key strength of this study design is the integration of a lived-experience consultant and structured opportunities for participant reflection. The study leadership team includes a consultant with lived experience of BPD who piloted all study procedures during setup and provided feedback on the overall research design. Participants are randomly identified to join focus groups and one-on-one qualitative interviews following completion of neurofeedback, gathering their reflections on the study experience and feedback on the intervention approach.

Embedding this element of assessment within the study design allows participants to share their perspectives in an open, low-pressure environment with an empathetic listener, without concern for meeting perceived researcher expectations. Importantly, the first round of interviews will occur before the R33 phase of the grant, allowing time to integrate participant feedback into the final study design.

### Risk Management

Although we anticipate our participants will generally be at low imminent risk for suicide based on our exclusion criteria, self-injurious behavior is a criterion for BPD (59). We therefore consider suicidal thoughts as behaviors will be expected in our patient population, and have consequently developed careful risk assessment and management procedures. Participants, when oriented to the study procedures, are alerted to the need for risk assessment throughout, as well as the circumstances under which confidentiality may be breached to assure participant safety, in compliance with law and ethical guidelines.

#### Safety plans

Once study eligibility is determined, participants collaborate with study staff to develop a safety plan following the Stanley Brown Safety Planning Intervention (60, 61). This plan aids participants in identifying personalized signs of an emerging crisis, specific strategies for managing distress, and resources and close others that they could contact in case of crisis. All participants are provided with a standardized set of local and national resources provided by the study team (988, Crisis Text Line, and local resources for New Haven or Boston depending on the participants’ location). A digital copy of the safety plan is e-mailed to participants directly upon completion of the planning meeting, and a physical copy is provided at scan day.

Participants are encouraged to view the safety plan as a living document, and are informed they can ask study staff to update it at any time. The plan remains accessible to study staff to support participants as-needed throughout the study. To ensure staff can respond appropriately in instances of crisis, we confirm participants’ home address by postcard and obtain information for <u>a</u> collateral contact who the participant agrees can be contacted by study staff if we lose contact with the participant during the study.

#### Risk assessment tool 1: UWRAP

Participants’ emotional state and urges for self-damaging behaviors are assessed routinely (on a 7-point scale) at baseline and at the end of each clinical assessment timepoint using the Linehan Risk Assessment Protocol (UWRAP) (62). Participants endorsing 4+ on any item are offered in-the-moment relaxation activities with study staff and coached to use coping strategies (e.g. contacting treatment providers, contacting close other).

#### Risk assessment tool 2: C-SSRS

We assess suicidal ideation and behavior at each clinical assessment timepoint (baseline, after NF, at the start of each DBT mindfulness module, and at follow-up months 1 and 2) using the C-SSRS.

#### Response to identified risk

If risk assessment tools or staff judgement raise concern for participant safety, a licensed study clinician is contacted for further assessment and safety planning. In particular, for any participant who has not yet developed a safety plan, endorses 4+ on any UWRAP item after engaging in suggested distress-management activities, or endorses type 4 or 5 ideation (suicidal ideation with intent or plan) or new suicidal behavior since last study visit on the CSSRS, a licensed clinician is immediately contacted for further assessment.

Endorsement of specific survey items indicating risk (i.e., item 9 on BDI) leads to an immediate programmed response in the survey software providing participants with their local crisis number, referring them to their personalized plans, and reminding them that survey responses are not reviewed in real time.

### Statistical Design and power analysis

#### Aims

In the R61 study phase, we aim to determine whether mbNF targeting the DMN modifies connectivity and improves mindfulness in participants with BPD, relative to control NF. The primary outcome is change in resting-state DMN connectivity from pre- to post-mbNF, either by decreasing within-DMN hyperconnectivity or by increasing DMN-CEN anticorrelation. The GO criterion for continuing to the R33 phase is greater pre- to post-NF change in at least one of these targets, with a medium effect size (d ≥ 0.5) between groups. The secondary outcome is change in self-reported mindfulness scores on the state mindfulness scale (SMS) from pre- to post-mbNF. We plan to randomize 52 participants (26 per group) in the R61 phase of the study. Because the outcome measures for this cohort are collected within two hours after randomization, little or no attrition is expected.

The R33 phase analyses focus on outcomes at later timepoints (after mbNF), over the course of the 20-week DBT skills group and two month follow-up period. This phase will assess the relationship between target engagement, clinical improvement, and longitudinal outcomes, including exploratory analyses to determine whether neural and symptom changes persist after mbNF and DBTsg. More participants (72, 36/group) are enrolled in this phase of the study because as much as 33% attrition is expected over the course of the full study.

#### Data preparation and statistics

Data will be examined for outliers, non-normal distributions, and non-linear associations. Preliminary analyses will examine potential covariates including site, age, sex, clinical variables, and head motion. Psychiatric service use (e.g., therapy, medication, unplanned mental health service use) will be assessed to test whether this differentially impacts (or is affected by) change in our neural targets. We will also explore baseline symptoms as a potential covariate, as differences in severity may lead to differences in target engagement. We will explore the presence of group effects (and time effects, when appropriate) across individual covariates or combined covariates using a liberal p <.05 uncorrected threshold and include any suprathreshold variable as a control covariate in subsequent analyses.

We will use a repeated measures ANOVA (or linear mixed-effects model in the presence of significant covariates) to test our hypothesis that mbNF will lead to larger decrease in DMN connectivity than control NF, with group (active vs. control) as a between-participant factor and time (pre-vs. post-NF) as the within-participant factor. This analysis will include all the R61 participants who complete the neurofeedback day (n = 26 per group). With a total sample size of 52 participants, assuming sphericity and a correlation of 0.5 among repeated measures, we will have 94% power (α=0.05) to detect a medium effect (Cohen’s *d*=0.50, Cohen’s *f*=0.25).

Missing data will be accommodated using multiple imputation or weighting procedures. Conclusions will primarily be based on effect sizes, with 95% confidence intervals, rather than statistical significance to maximize reproducibility. Based on our prior studies, we estimate ∼30% attrition/data loss and have planned recruitment to support data collection on primary and secondary outcomes from 52 participants.

#### Data management, sharing, and monitoring

Data are stored in HIPAA-compliant servers (MR data), REDCap databases (clinical and self-report data) and TEAMS folders (interview recordings). In accordance with funder guidelines, we will share data through the National Data Archive (NDA) by assigning each participant a global user identifier and uploading data using NDA standards.

#### Status and Timeline

The study enrolled the first participant in May 2025. We anticipate that R61 phase recruitment will be complete in summer 2026 with results expected shortly thereafter.

## Discussion

BPD is a persistent and severe psychiatric disorder that results in missed social and occupational opportunities in early adulthood and increases risk for suicide. There is an urgent need to improve the efficacy of treatments and the durability of their effects. This R61-R33 project tests a novel intervention approach that targets core mechanisms underlying BPD. Specifically, mindfulness with mbNF, a noninvasive, personalized therapeutic tool, is designed to normalize connectivity of the DMN as well as reduce BPD symptoms.

If successful in improving clinical outcomes for adults with BPD, we will be positioned to a) develop methods to identify patients most likely to respond to mbNF+DBTsg and to develop a better understanding of mbNF dosage and timing considerations, and b) to expand our understanding of downstream and larger-scale neural circuit changes that occur in response to mbNF alone and in combination with DBTsg. Further expanding on this work, we aim to determine when and how to use mbNF to optimize symptom reduction and mindfulness utilization. For example, for participants who experience decreased mindfulness utilization or recurrence of clinical symptoms post-treatment, “mbNF booster sessions” could be administered. This personalized treatment approach is directly in line with precision medicine initiatives, and ultimately, would revolutionize clinical care for people with BPD.

While this project zeroes in on DMN and CEN, we recognize that these networks reside within larger neural systems relevant for understanding the mechanisms of both pathology and treatment response in BPD. Given the extensive imaging data correlating amygdala dysregulation with BPD symptomatology (63, 64), and evidence from neuromodulation research that altering frontal/executive networks has downstream effects on limbic networks e.g. (65), the proposed project will open the door to more precisely articulating the relevant network-level mechanisms.

## Data Availability

No datasets were generated or analysed during the current study. All relevant data from this study will be made available upon study completion.

## Acknowledgements

We would like to acknowledge the staff who have joined our study team to make this project’s daily operations possible, and the important contributions of the study participants.

## Author contributions

Katherine G. Jones: Writing: original and revised drafts, protocol design and implementation, figure design

Marlee M. Vandewouw: Writing: original and revised drafts, protocol design and implementation, fMRI analysis design and implementation

Jitendra Awasthi: Writing: revised drafts, protocol implementation, fMRI analysis design and implementation, fMRI neurofeedback system implementation

Alexandra A. Alario: Writing: revised drafts, protocol implementation

Clemens Bauer: mindfulness-based fMRI neurofeedback initial design and adaptation for this protocol, funding acquisition, writing: revised drafts

Beth Brewer: protocol implementation, writing: revised drafts Nicole Campbell: protocol implementation, writing: revised drafts Dominic Denning: protocol implementation

Keara D. Greene: protocol implementation, team coordination and central administration, writing: revised drafts

Jude Hammoud: protocol implementation, writing: revised drafts

Oliver Hinds: MURFI neurofeedback software initial design and adaptation for this protocol Sarah Huffman: protocol implementation

Connie Maerker: protocol implementation Sarah Paprotna: protocol implementation

Maolin Qiu: fMRI and MURFI neurofeedback adaptation and setup for this protocol Benjamin Swinchoski: protocol implementation, writing: revised drafts

Anna Taylor: protocol implementation Elinor Waite: protocol implementation

Paul Wighton: fMRI and MURFI neurofeedback adaptation and setup for this protocol Maya Whaley: protocol implementation

Jillian Papa: funding acquisition, writing: revised drafts, protocol implementation, design and implementation of program evaluation

Katherine Dixon Gordon: funding acquisition, writing: revised draft, protocol implementation, design, implementation and supervision of psychotherapy procedures

Michelle Hampson: funding acquisition, writing: revised draft, protocol implementation, supervision of fMRI and MURFI and fMRI analysis implementation

Susan Whitfield-Gabrieli: initial design and implementation of mindfulness-based fMRI neurofeedback and MURFI neurofeedback system, protocol design and funding acquisition, supervision of protocol implementation, writing: revised draft

Sarah K. Fineberg: funding acquisition, protocol design, supervision of overall team for implementation, close supervision of clinical and risk management procedures, writing: initial and revised drafts

---

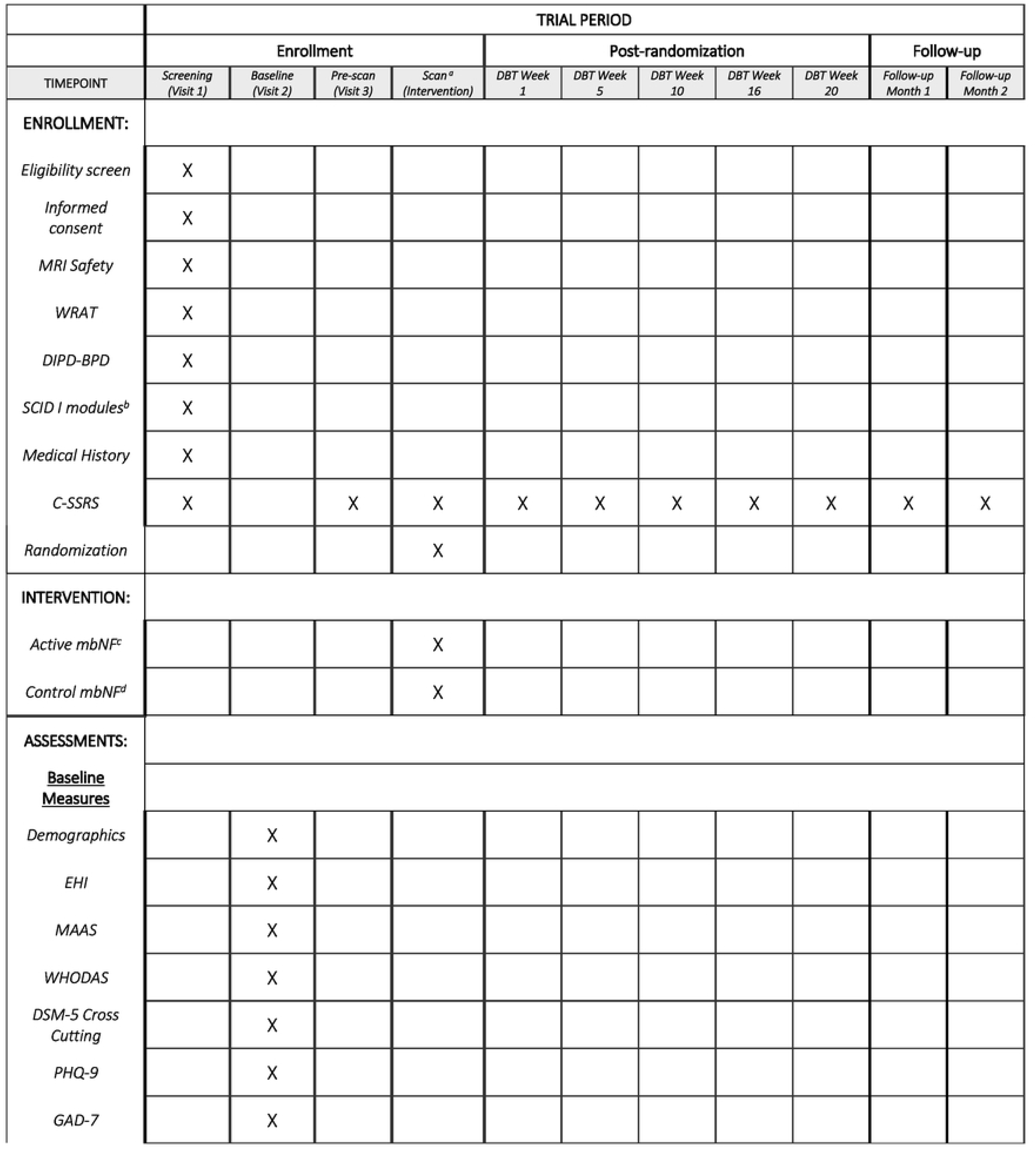
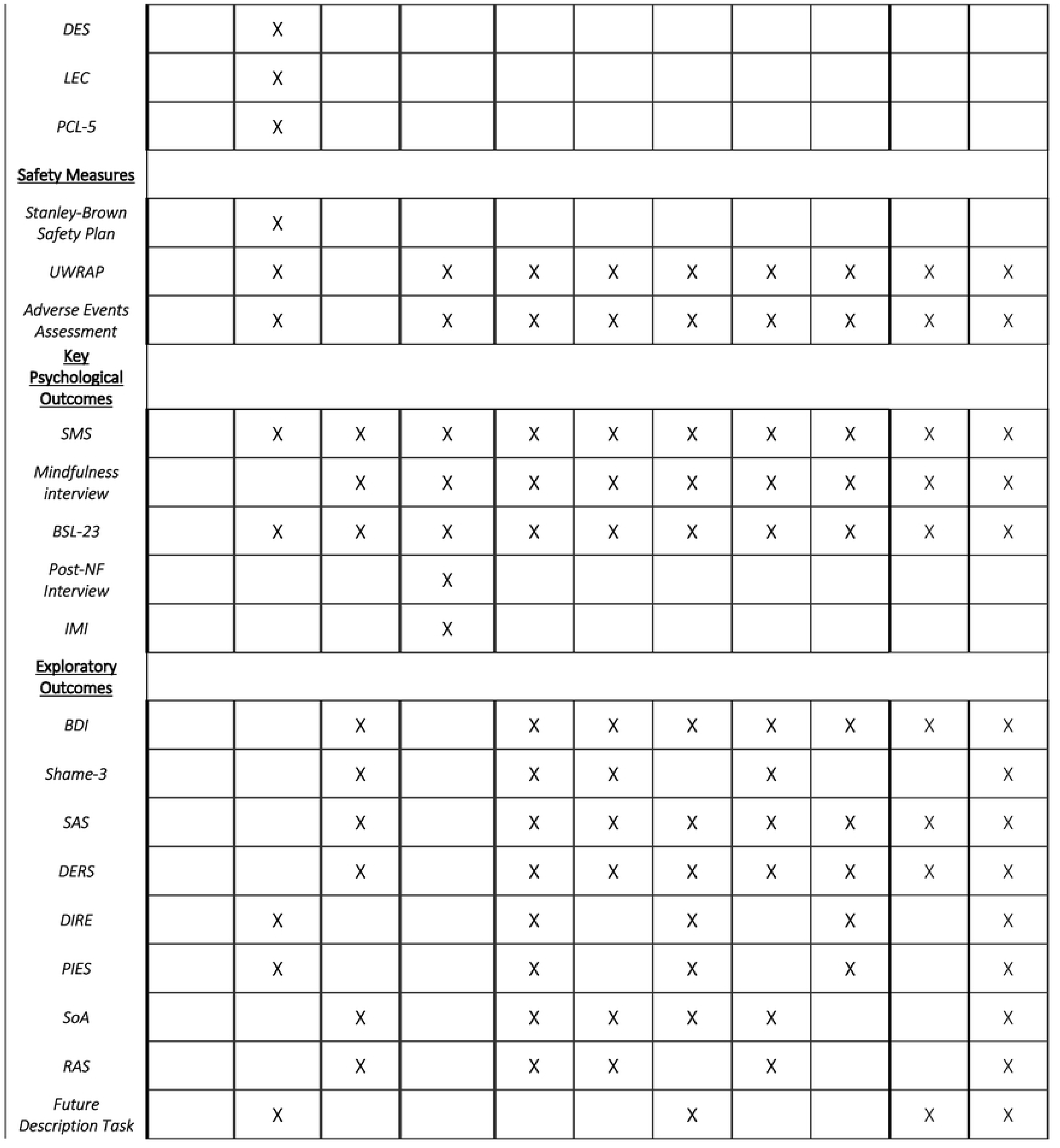
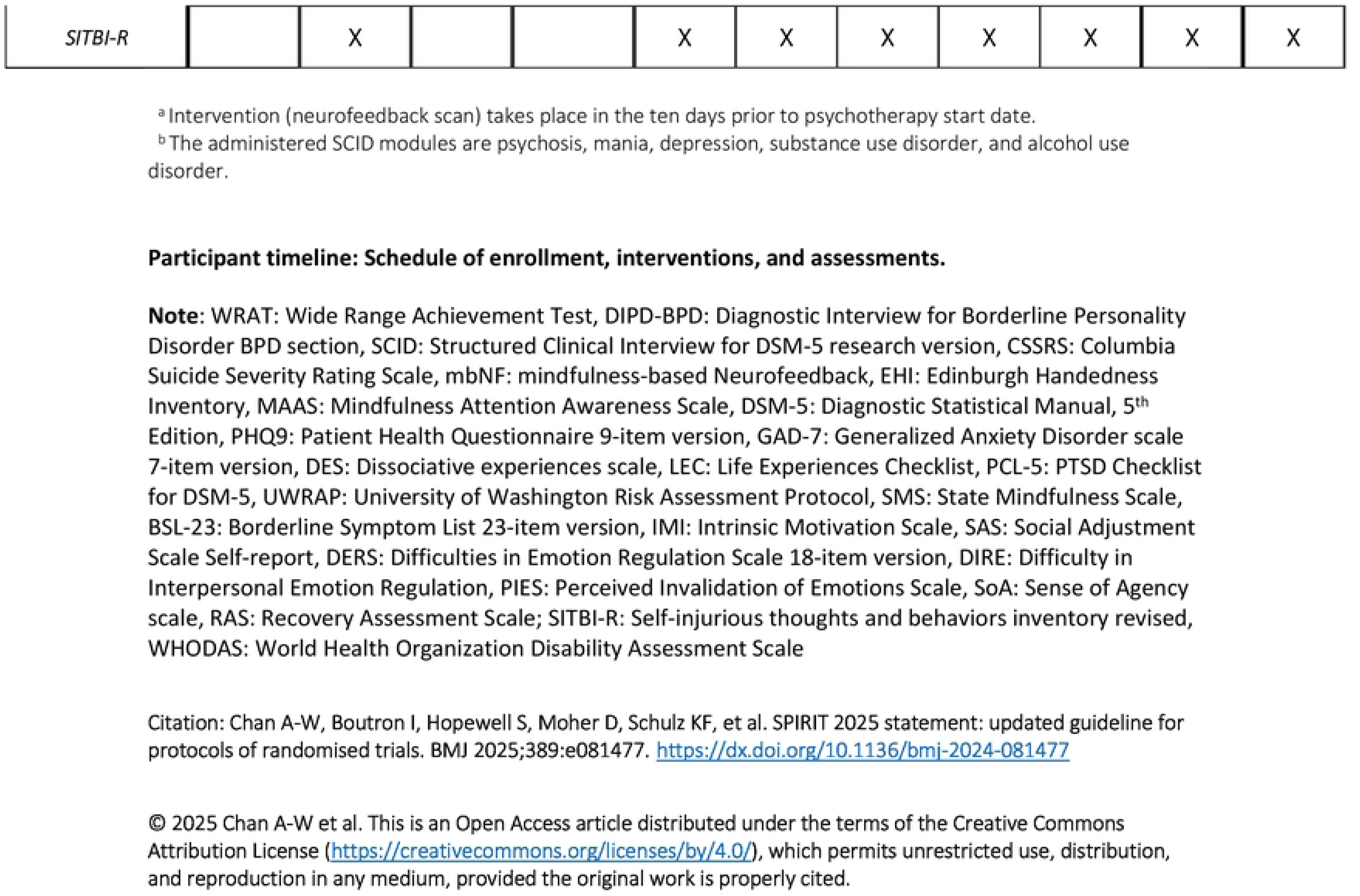

